# Diagnostic accuracy study of a novel blood-based assay^#^ for identification of TB in people living with HIV

**DOI:** 10.1101/2020.06.10.20127209

**Authors:** Erik Södersten, Stefano Ongarello, Anna Mantsoki, Romain Wyss, David H. Persing, Sara Banderby, Linda Strömqvist Meuzelaar, Jacqueline Prieto, Devasena Gnanashanmugam, Purvesh Khatri, Samuel G. Schumacher, Claudia M. Denkinger

**Author notes:** Contributed equally (Samuel G. Schumacher, Claudia M. Denkinger). Disclaimer: Product in development. Not for use in diagnostic procedures. Not reviewed by any regulatory body.

## Abstract

A non-sputum triage test to rule out TB disease is a WHO high-priority diagnostic and a combinatory score based on a 3-gene host-signature has shown promise in discriminating TB from other illnesses. We evaluated the accuracy of an early-prototype cartridge-assay (“Xpert MTB Host Response”, or Xpert-MTB-HR-Prototype) of this 3-gene signature on bio-banked blood-samples from PLHIV against a comprehensive microbiological reference standard (CMRS) and against Xpert® MTB/RIF on first sputum alone. We depict results based on performance targets set by WHO in comparison with a laboratory-based CRP assay. Of 201 patients included, 67 were culture-positive for *Mycobacterium tuberculosis*. AUC for the Xpert-MTB-HR-Prototype was 0·89 (CI 0·83-0·94) against the CMRS and 0·94 (CI 0·89-0·98) against Xpert MTB/RIF. Considering Xpert-MTB-HR-Prototype as a triage test (at nearest upper value of sensitivity to 90%), specificity was 55·8% (CI 47·2-64·1) compared to the CMRS and 85·9% (CI 79·3-90·7) compared to Xpert MTB/RIF as confirmatory tests. Considering Xpert-MTB-HR-Prototype as a stand-alone diagnostic test, at a specificity near 95%, the test achieved a sensitivity of 65·7% (CI 53·7-75·9) while CRP achieved a sensitivity of only 13·6% (CI 7·3-23·4). In this first accuracy study of a prototype blood-based host-marker assay, we show the possible value of the assay for triage and diagnosis in PLHIV.

**One Sentence Summary:** This is an accuracy study of a blood-based host-marker assay demonstrating its potential in triage and diagnosis of active tuberculosis (TB) in people living with HIV (PLHIV).

## Introduction

In 2018, 10 million people developed tuberculosis disease (TB) and 1·5 million people lost their lives to the illness *(1)*. According to the WHO’s “End TB Strategy”, improved diagnostic tests are required as a means to stop the global tuberculosis epidemic by 2035. The prioritized diagnostic tests to be developed were defined in a target setting exercise in 2014 (*1*). One of the highest priority target product profiles (TPPs) is a non-sputum triage test to rule out disease with a minimum of 90% sensitivity and 70% specificity (optimally 95% sensitivity and 80% specificity) to be used by first-contact providers to identify patients who need further confirmatory testing (*1*). Currently no triage test can be done to meet these targets from a non-sputum sample. Chest X-ray is widely used as a screening test, but is limited by the infrastructure and instrumentation needs and its lack of specificity for TB (*2*). An optimal novel test should be performable on a readily accessible patient sample, such as blood or urine and have limited infrastructure needs (*1, 3*). Detection of C-reactive protein (CRP) on lateral-flow-tests is currently the only test that meets operational characteristics for a triage test, but performance evaluations show limited specificity and have been mostly restricted to populations with high HIV prevalence (*4*).

However, novel tests show promise. In 2016, Sweeney et al published a multi-cohort study based on 14 publicly available microarray data sets obtained from whole-blood samples from patients with symptoms compatible with TB disease. The study suggested that a combinatory score (TB-score) based on blood mRNA levels of only three differentially expressed genes (GBP5, DUSP3 and KLF2) can discriminate between TB disease and other diseases (*5*).

Cepheid (Sunnyvale, CA, USA) has developed an early prototype GeneXpert PCR test, Xpert-MTB-HR-Prototype, that quantifies relative mRNA-levels of the 3-gene signature in a patient whole-blood sample. The Xpert-MTB-HR-Prototype test is intended for direct analysis of fingerprick blood but can in addition be utilized retrospectively with blood samples that have been conserved in PAXgene buffer. This study is a first feasibility study to evaluate the performance of Cepheid’s novel assay prototype on preserved blood samples from PLHIV. PLHIV are a particularly vulnerable patient group with high proportions of extra-pulmonary TB or cases of pulmonary TB that are unable to produce sputum (*6*).

Based on the data provided by Sweeney et al both a triage and a stand-alone diagnosis use-cases for the signature can be envisioned. Here, we assess the performance of the novel blood-based 3-gene signature cartridge prototype as a non-sputum-based triage test or a stand-alone diagnostic test for TB disease against a microbiological reference standard (culture and Xpert® MTB/RIF) and separately, which is more likely to occur when implemented in limited-resource settings and thus recommended by the World Health Organization (WHO), against Xpert MTB/RIF alone.

## Results

### Subhead 1: Performance evaluation of the Xpert-MTB-HR-Prototype against the comprehensive microbiological reference standard

A total of 201 samples from a South African and Peruvian cohort of patients diagnosed with HIV were included in the study. Of the 201 patients, 67 (33·3%) were diagnosed with TB with 23 (34·3%) being smear negative/culture positive and 44 (65·7%) being smear positive/culture positive). Five patients were defined as “Subclinical TB” and 129 (64·2%) were categorized as “Non-TB” patients (46 LTBI, 83 non LTBI). No patient with possible TB was included. Xpert MTB/RIF on the first sputum sample was able to identify 53 patients (79·1%; smear positive 43/44 [97·7%] and smear negative 10/23 [43·5%]), while Xpert MTB/RIF from all sputum samples detected 57 patients (85%).

The patient characteristics are reported in Table 1. The median age was 36 with 65% of participants being female. All patients had confirmed HIV infection and for all but 10 the CD4 count was available, with a median CD4 at 375 and 58 patients with a CD4 <200 cells/mm^3^. All but 20 patients had more than three symptoms suggestive of TB.

**Table 1:** Patient characteristics.

| Patient Characteristics |  | N (total = 201) |
| --- | --- | --- |
| **Age** | Median (IQR) | 36 (31-43) |
| **Sex** | Female (%) | 130 (65·0) |
|  | Male (%) | 71 (35·0) |
| **TB status** | TB+ S-C+ (% of culture positive) | 23 (34·3) |
|  | S+C+ | 44 (65·7) |
|  | TB – (% of all) | 134 (66·7) |
| **CD4 Count (cells/mm<sup>3</sup>)** | <200 (% with CD4 count) | 58 (30·0) |
|  | ≥200 (% with CD4 count) | 133 (70·0) |
|  | Unknown (% of total) | 10 (5·0) |
|  | Median (IQR) | 375 (154 – 596) |
| **History of BCG** | Positive history of vaccination (% of total) | 177 (88·1) |
|  | Negative history of vaccination (% of total) | 4 (2·0) |
|  | Unknown history of vaccination and scar indeterminate (% of total) | 20 (10·0) |
| **Prior history of TB** | Positive (% of total) | 128 (63·7) |
|  | Negative (% of total) | 73 (36·3) |
| **QuantiFERON result** | Positive (% with result) | 78 (38·8) |
|  | Negative (% with result) | 95 (47·3) |
|  | Indeterminate (% with result) | 26 (12·9) |
|  | Not obtained (% of total) | 2 (1·0) |
| **Site of Study** | South Africa | 195 (97·0) |
|  | Peru | 6 (3·0) |

The TB-score values across participants with and without TB as defined by CMRS, further subdivided by Xpert MTB/RIF result on first sputum, are depicted in Figure 1(A). Figure 1(B) shows the same for CRP. Interestingly, 8 patients identified by CMRS with paucibacillary diseases were negative on both TB-score and CRP.

**Figure 1:**
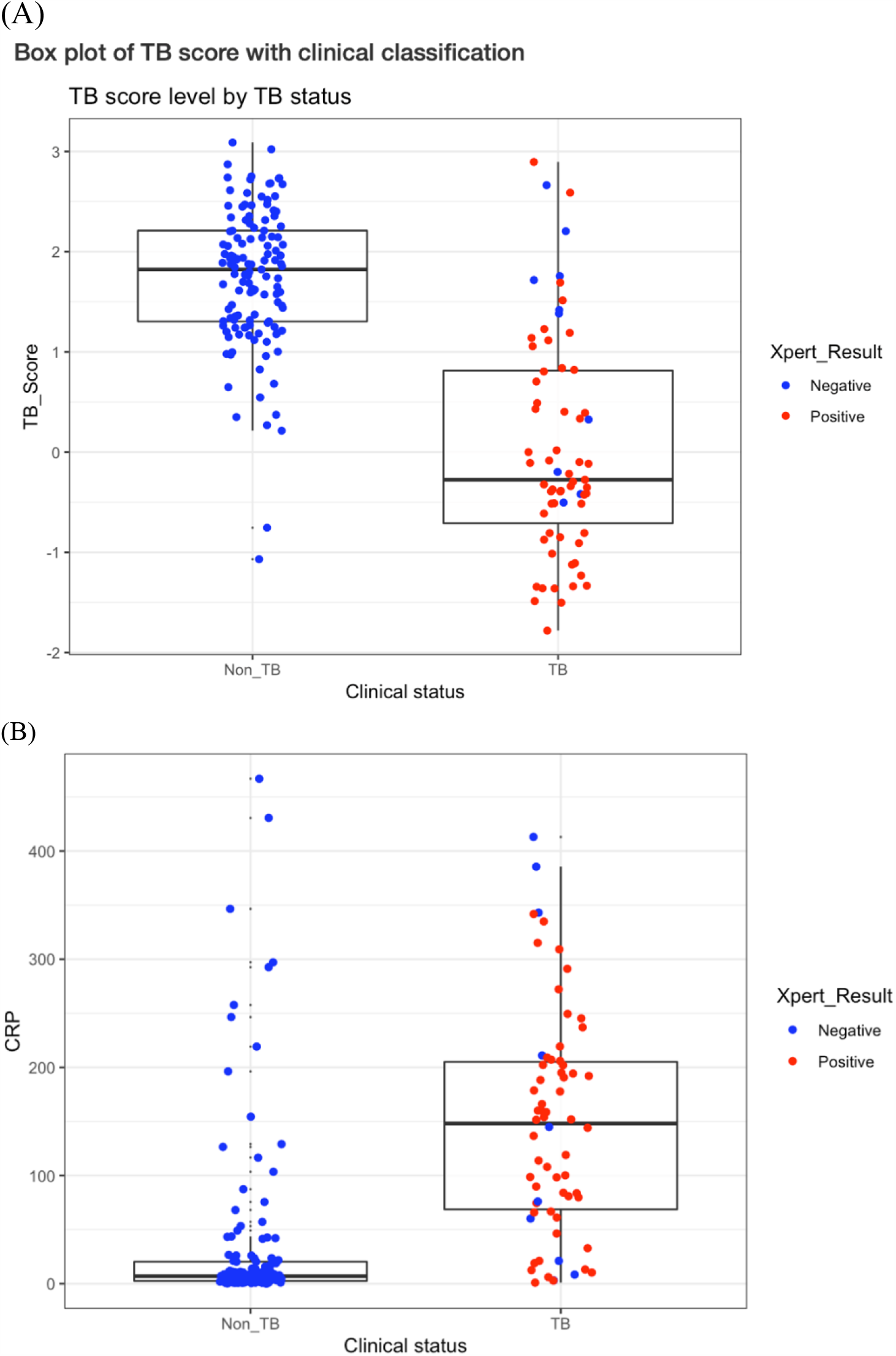
Values for (A) Xpert-MTB-HR-Prototype and (B) laboratory-based CRP against a comprehensive microbiological reference standard subdivided by Xpert MTB/RIF result on first sputum.

The performance of the Xpert-MTB-HR-Prototype was first evaluated against the CMRS which yielded an AUC of = 0·89 (CI 0·83-0·94) (Figure 2). As comparison, CRP yielded an AUC = 0·86 (CI 0·80-0·91) (Figure 2). Using DeLong’s test the AUROCs for the Xpert-MTB-HR-Prototype and CRP were not statistically different (p>0.3) (*7*). Optimal sensitivity and specificity were obtained for the Xpert-MTB-HR-Prototype when the Youden index was maximized. At this TB-score cut-point sensitivity was 77·6% (95% Confidence Interval [CI] 66·3-85·9), specificity was 92·2% (CI 86·3-95·7) (Figure 2). In comparison, the CRP at the Youden index was 60·3mg/L, which translated into 80·3% sensitivity (CI 69·1-88·1) and 86·8% specificity (CI 79·9-91·6; Figure 2).

**Figure 2:**
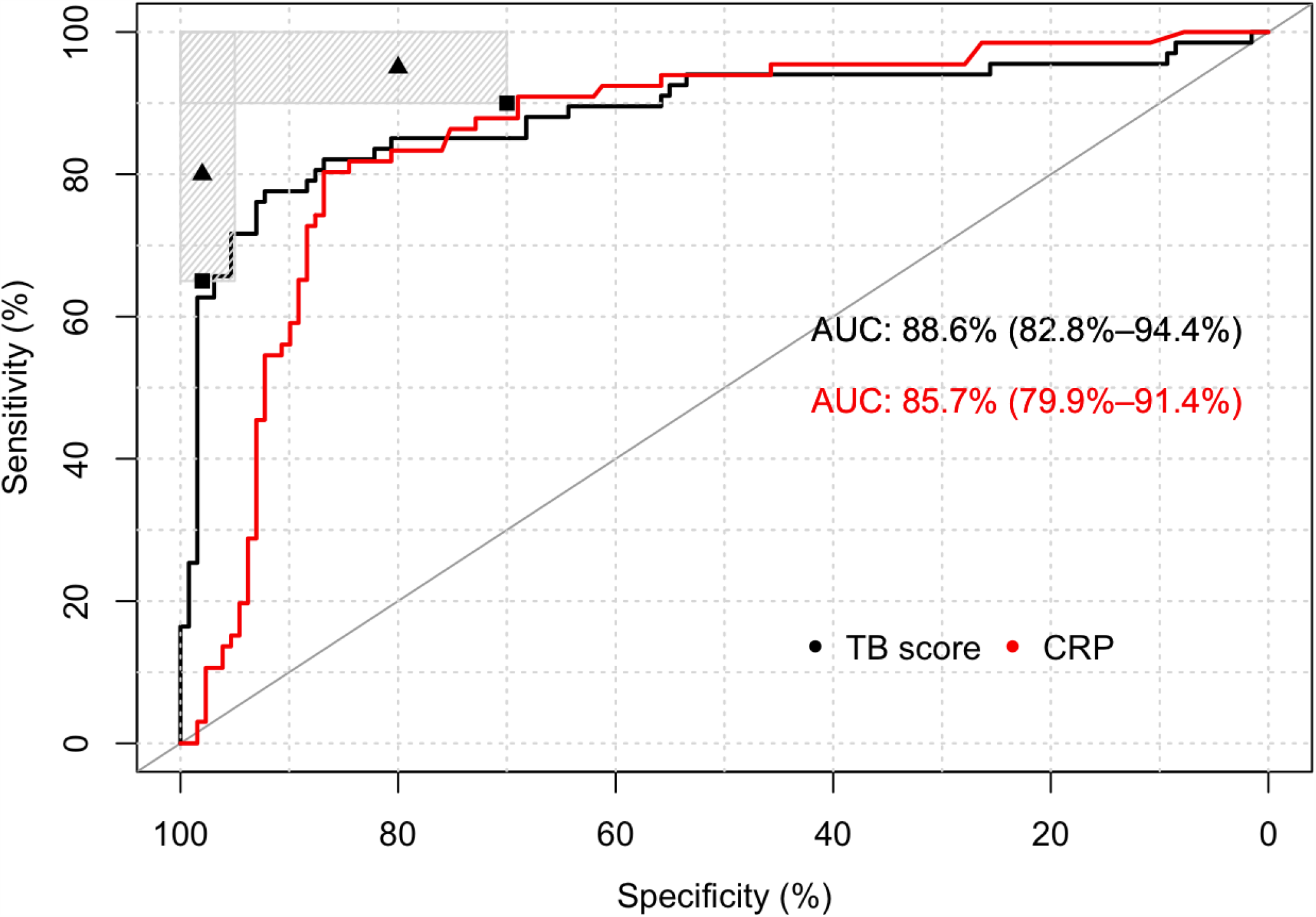
ROC curve for Xpert-MTB-HR-Prototype test and laboratory-based CRP test against a comprehensive microbiological reference standard. Caption: The shaded regions represent areas with sensitivity and specificity combinations that meet at least the minimal target of one of the TPPs (triage or non-sputum diagnostic). The triangles represent the optimal, the square the minimal targets. The black line represents the TB-score, the red line the CRP.

Considering the Xpert-MTB-HR-Prototype as triage test at fixed value of sensitivity near 90% (91·0% nearest upper value; CI 81·8-95·8), the corresponding specificity was 55·8% (CI 47·2-64·1) against a CMRS. At fixed value of sensitivity near 95% (95·5% nearest upper value; CI 87·6-98·5), the specificity was 25·6% (CI 18·8-33·7). In comparison, the laboratory-based CRP test at fixed value of sensitivity near 90% (90·9% nearest upper value; CI 81·6-95·8), would achieve a specificity of 69·0% (CI 60·6-76·3; CRP-value of 12·3 mg/L) against CMRS. And similarly, at a fixed value of sensitivity near 95% (95·5% nearest upper value; CI 87·5-98·4), the specificity was 45·7% (CI 37·4-54·3) (CRP-value of 6·2mg/L).

Considering the Xpert-MTB-HR-Prototype test as a stand-alone diagnostic test, specificity should ideally be at least 95% against CMRS. At a specificity of 95·3% (nearest value; CI 90·2-97·9), the test achieves a sensitivity of 65·7% (CI 53·7-75·9). In comparison, the CRP-test performance at a specificity of 95·4% (nearest value; CI 90·2-97·9), resulted in a sensitivity of only 13·6% (CI 7·3-23·4) (CRP-value of 253·6 mg/L).

### Subhead 2: Performance evaluation of the Xpert-MTB-HR-Prototype against Xpert MTB/RIF

The TPP of the WHO specified a minimal sensitivity and specificity for a non-sputum-based triage test against Xpert MTB/RIF as the most likely confirmatory test in high-burden, limited resource settings. Therefore, we assessed the performance against a single Xpert MTB/RIF on the first sputum sample. AUC for the Xpert-MTB-HR-Prototype with this reference standard was 0·94 (CI 0·89-0·98) whereas AUC for CRP was 0·86 (CI 0·80-0·91) (Figure 3). Using DeLong’s test the AUROC for the Xpert-MTB-HR-Prototype was significantly higher than that of CRP (p=3.85e-3) (*7*). Optimized to achieve the highest Youden index, the Xpert-MTB-HR-Prototype had 88·7% (CI 77·4 – 94·7) sensitivity, 89·4% (CI 83·3 – 93·5) specificity) (Figure 3). Considering the Xpert-MTB-HR-Prototype test as a triage test at fixed value of sensitivity near 90% (90·6% nearest upper value; CI 79·8-95·9), the Xpert-MTB-HR-Prototype specificity was 85·9% (CI 79·3-90·7). At fixed value of sensitivity near 95% (96·2% nearest upper value; CI 87·3, 99·0), the specificity was 78·2% (CI 70·7, 84·2).

**Figure 3:**
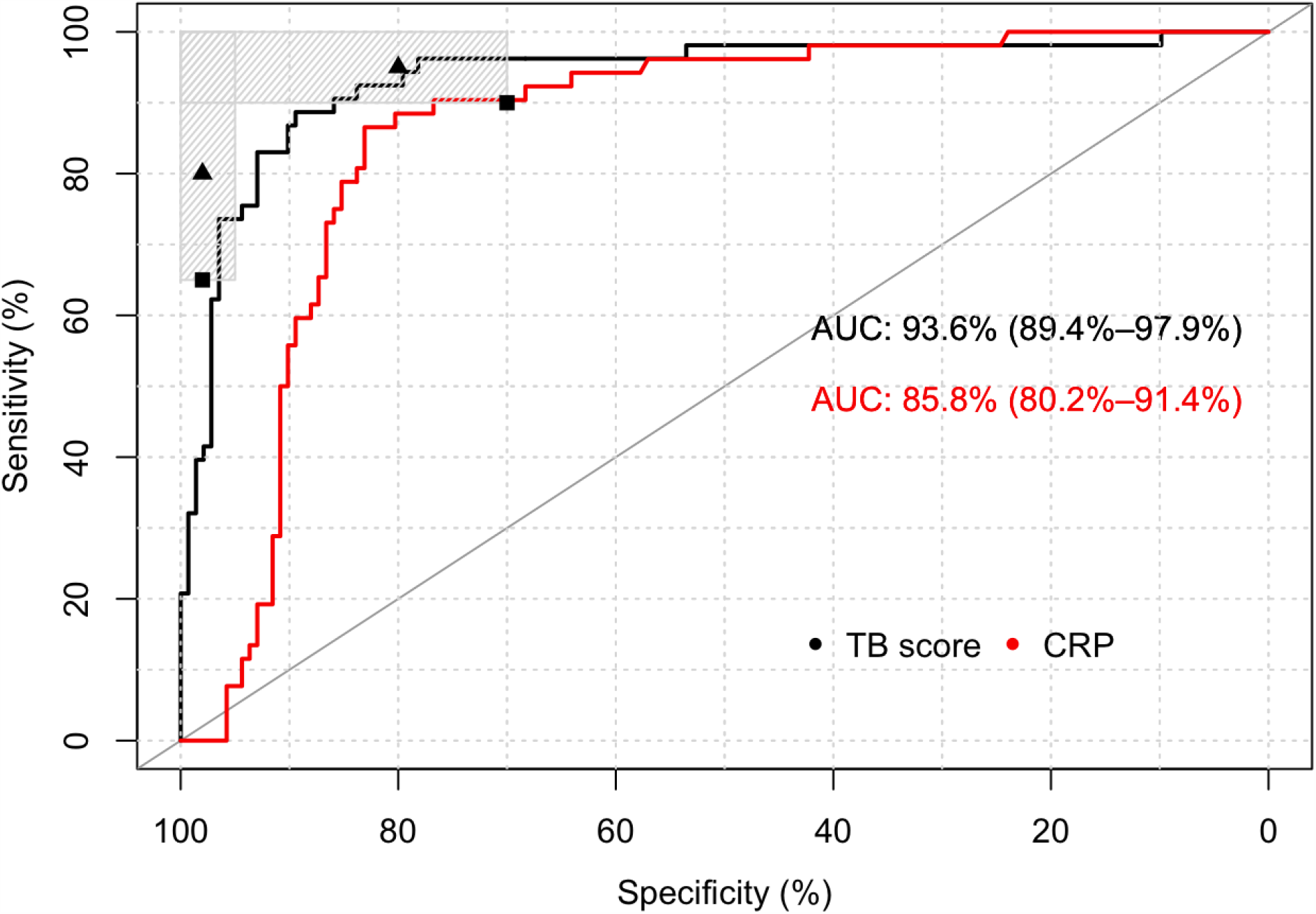
ROC curve against Xpert MTB/RIF on first sputum as reference standard for Xpert-MTB-HR-Prototype test and laboratory-based CRP test. Caption: The shaded regions represent areas with sensitivity and specificity combinations that meet at least the minimal target of one of the TPPs (triage or non-sputum diagnostic). The triangles represent the optimal, the square the minimal targets. The black line represents the TB-score, the red line the CRP.

In comparison, at the CRP test value of 60·3 mg/L (optimized to achieve the highest Youden index against the Xpert MTB/RIF reference standard), sensitivity was 86·5 (CI 74·7 – 93·3) and specificity was 82·4 (CI 75·3-87·8). At fixed value of sensitivity near 90% (90·4% nearest upper value; CI 79·4-95·9), achieved a specificity of 76·8% (CI 69·2-82·3) (CRP-value of 29·7mg/L). And at a sensitivity of near 95% (96·2% nearest upper value; CI 87·0-98·4), the specificity was 57·0% (CI 48·9-64·9) (CRP-value of 10·2mg/L) (Figure 3).

When comparing Xpert-MTB-HR-Prototype and CRP scores for each sample, we observed that the samples misclassified by Xpert-MTB-HR-Prototype and CRP are largely the same (see Supplement Figure 2; 4^th^ quadrant).

Subgroups analyses of sensitivity and specificity at the optimal TB-score cut-point by CD4 count and by number of symptoms for the Xpert-MTB-HR-Prototype are provided in Table 2 (ROC curve and box plots for subgroups are in Supplement Figures 4 and 5). Subgroup analysis assessing specificity according to LTBI status as defined by IGRA, determined a lower specificity in patients with LTBI 89·1% (CI 77·0-95·3) versus 94·0% (CI 86·7-97·4) but confidence intervals were overlapping (box plots by LTBI status in Supplement Figure 6). In the participants identified to have “Subclinical TB”, 0 out of 5, had a positive Xpert-MTB-HR-Prototype test, when the optimized cut-point was used.

**Table 2:** Xpert-MTB-HR-Prototype subgroups analyses at optimal TB-score cut-point against Xpert MTB/RIF on first sample as reference standard.

| SUBGROUP | N | SENSITIVITY % (CI) | SPECIFICITY % (CI) |
| --- | --- | --- | --- |
| **CD4** |  |  |  |
| ≥200 | 129 | 75.0 (50.5 – 89.9) | 94.7 (88.9 – 97.5) |
| <200 | 56 | 93.8 (79.9 – 98.3) | 66.7 (46.7 – 82.0) |
| **SYMPTOMS** |  |  |  |
| ≤3 | 19 | 100 (70.1 - 100) | 80.0 (49.0 – 94.3) |
| >3 | 176 | 86.4 (73.3 – 93.6) | 90.2 (83.9 – 94.2) |
| **LTBI (IN NON TB)** |  |  |  |
| NON-LTBI | 82 | - | 93.9 (86.5-97.4) |
| LTBI | 46 | - | 89.1 (77.0-95.3) |

Abbreviations: CI – confidence interval; LTBI – latent tuberculosis infection

An exploratory analysis assessed time to culture positivity against the TB-score (Supplement Figure 3). From this analysis, it was evident that many of those cases not captured by set cut-points of sensitivity at 90% or 95% were those with paucibacillary disease, i.e. long time to culture positivity, with many also not captured by Xpert MTB/RIF (Supplement Figure 2).

Considering the implementation of the Xpert-MTB-HR-Prototype as a triage test at a sensitivity of 90·6% (specificity 85·9%) followed by an Xpert MTB/RIF as a confirmatory test, 68 out of 196 patients (excluding 5 subclinical cases) would have a positive result with the triage test (48 true positive and 20 false positive cases) and would have to be confirmed by Xpert MTB/RIF on the first sputum. The algorithm of Xpert-MTB-HR-Prototype followed by Xpert MTB/RIF would miss 19 cases identified by CMRS compared to 14 cases missed with a strategy of Xpert MTB/RIF alone on all presumed TB patients. This strategy would utilize 196 Xpert-MTB-HR-Prototype cartridges and 69 Xpert MTB/RIF cartridges. In comparison the CRP-test (at a sensitivity of 90·4%; specificity 76·8%), would miss the same number of cases but utilize 81 Xpert MTB/RIF cartridges in addition to the 196 CRP tests.

## Discussion

This study assesses for the first time the performance of a novel, blood-based early prototype test that is based on the quantification of mRNA-levels of 3 host genes in well characterized samples from PLHIV presenting with symptoms suggestive of TB in high endemic settings. The study establishes the feasibility of implementing the 3-gene signature in a well-proven cartridge-based testing system. Furthermore, the study confirms the performance of the signature in this cartridge system in comparison to various studies with RNA sequencing and qPCR and again shows its ability to meet the minimal WHO TPP for a triage test (*8*). The signature that was implemented in the cartridge is the most extensively validated transcriptomic signature to date, including a validation by an independent group in a large cohort of >1400 patients (*9*). It was originally derived through meta-analysis of whole-blood gene-expression data of 3 clinical cohorts from 6 different countries, and subsequently validated in datasets from 11 countries, in which it distinguished active TB disease from other diseases with sensitivity and specificity estimates of 81% and 74%, respectively (*8*). Importantly, the 3-gene signature was also shown to predict progression from LTBI to active TB up to 6 months prior and associated with risk of subclinical active TB post-treatment in independent cohorts (*10*).

Our study shows that in HIV-positive patients with active TB the performance of the Xpert-MTB-HR Prototype as a triage test is similar to that of a laboratory-based CRP assay at pre-selected 90% sensitivity compared to CMRS. However, the TPP for a non-sputum-based triage test by the WHO is specified against Xpert MTB/RIF as the most likely confirmatory test in high-burden, limited resource settings. When using Xpert MTB/RIF as the confirmatory test on the first sputum in a triage scenario, the prototype cartridge was significantly better than CRP. Using Xpert MTB/RIF on the first sputum as a reference standard at the pre-selected sensitivity of 95% (i.e. optimal target), the Xpert-MTB-HR-Prototype achieved 78% specificity, which approaches the optimal TPP for a non-sputum triage test specified by the WHO. In contrast, at 95% sensitivity, laboratory-based CRP assay has substantially lower specificity (57%).

This significantly higher accuracy of the Xpert MTB-HR prototype compared to CRP has important implications for a triage test, especially in a HIV-negative patient population. In HIV-negative cohorts, performance of CRP is highly likely more affected by non-TB inflammatory conditions. CRP reflects ongoing inflammation and/or tissue damage across a broad range of diseases including infections, allergic complications of infection, autoimmune diseases and malignancies such as lymphoma, carcinoma, and sarcoma. Examples of routine clinical uses of CRP include (1) assessment of disease activity in inflammatory conditions (e.g., rheumatoid arthritis, Crohn’s disease, psoriatic arthropathy, rheumatic fever, acute pancreatitis, familial fevers), (2) diagnosis and management of infection such as bacterial endocarditis, intercurrent infection in lupus and leukemia, and (3) predicting future atherothrombotic events, including coronary events, stroke, and progression of peripheral arterial disease (*11*). When used in HIV-negative population for triage, where other inflammatory conditions are more prevalent, specificity of CRP will further reduce, and result in higher number of false positives requiring additional confirmatory tests. In contrast, performance for the 3-gene signature in other studies was shown to be lower in HIV-positive patients with TB than in HIV-negative patients, which might relate to the lesser immune-stimulation in HIV-positive patients (*5*). Hence, the Xpert-MTB-HR-Prototype can be expected to have higher accuracy in larger cohort of those without HIV and may be able to achieve the optimal TPP for a non-sputum triage test.

At 90% sensitivity compared to CMRS, as expected, both the prototype and CRP missed 6 patients. The median (and average) CD4 count for the 6 patients missed in both tests is 524. These results suggest that in PLHIV some patients may not mount sufficient/detectable host response as represented by either Xpert-MTB-HR-Prototype or CRP, even though they are not substantially immunosuppressed. The paucibacillary nature of the disease in these patients might be contributing (all but one of these patients were smear-negative) to the limited immune stimulation. Collectively, our results suggest that the 10% of patients with ATB missed by the prototype cartridge are those with limited host response due to paucibacillary infection that may lead to longer time to positivity.

Thus, a larger cohort including HIV-positive and HIV-negative patients should be explored to assess how the two tests perform in those subgroups in comparison. Also, the current CRP results were generated with a laboratory-based platform (Abbott Architect), while point-of-care CRP tests are available and most suitable for use as a triage test (*4*). Thus, a comparison of the Xpert-MTB-HR-Prototype in comparison to a point-of-care CRP would be a logical next step and an important study in real world application of both tests to assess whether the results observed here in comparison to CRP hold up when a simple lateral-flow assay for CRP is used.

The results of the Xpert-MTB-HR-Prototype test as a stand-alone diagnostic test seen in our study, hold promise particularly for patients who are unable to provide a sputum sample, e.g. PLHIV and children. These data also need to be further validated and studies to assess the early prototype for triage testing in children are ongoing. Considering the increased use of the Xpert MTB/RIF Ultra assay, future studies will need to include comparisons using Ultra to understand the role of the Xpert-MTB-HR-Prototype in these settings. The Sweeney et al study also suggested that the 3-gene signature may be useful for monitoring the response to TB treatment as the TB-score normalizes in patients with successful treatment and studies to evaluate the Xpert-MTB-HR-Prototype for treatment monitoring are also ongoing (*5*). Given that we tested an early prototype of the cartridge, further improvements in its diagnostic performance are conceivable with refinements in its development.

Although these data hold promise for the performance of the Xpert-MTB-HR-Prototype, widespread access and use will depend on costs and programs that enable access to the cartridge. Widespread use is unlikely to occur if costs are much greater than currently available Xpert MTB/RIF tests. Current pricing systems for other Xpert products in countries with a high burden of tuberculosis are likely to be employed when this product is available to the market.

The study has several limitations. Firstly, all samples used in the study were from HIV-positive subjects. In active screening, large proportion of subjects are expected to be HIV-negative, where the 3-gene TB signature has been shown to have higher accuracy. Secondly, while this was a nested case-control study, the degree of bias is probably very limited, as controls were selected from the same population as the cases and the characteristics of the population in terms of TB prevalence and smear status are similar to that of the total cohort. Thirdly, as the study was used as a derivation cohort to select an optimal cut-point and use-case, the performance needs to be further evaluated in a validation cohort with a set cut-point. Fourthly, the study only assessed samples from South Africa and Peru and sample size from Peru was too small to perform a subgroup analysis by country of origin. While South Africa contributes a significant share of the global TB/HIV epidemic, it will be critical to validate the host-marker based signature in other countries where there are more co-infections likely (e.g. in equatorial Africa), other strains circulating and other host genetic backgrounds. However, as we noted above, the 3-gene TB signature itself has been extensively validated across different countries; thus, substantially reducing the likelihood of effects of other circulating strains and host genetics on its performance. Lastly, another limitation is that testing of the samples was performed non-blinded to the results of the reference standard by the test manufacturer. However, given that this was the first accuracy study on the cartridge, and no preset cut-point was available, blinding was not possible as a cut-point needed to be defined. Given that no subjective interpretation is necessary of the test, this is not likely to introduce bias. And importantly, all data was made available to analyze independently to FIND.

The study also has several strengths. The protocol and analysis plan were predefined. The assessment was done in a cohort that presented with symptoms suggestive of TB in a high-prevalence setting. The controls were chosen from the same cohort as the cases. The prevalence and smear-status of the cohort are similar to what has been reported in other TB accuracy studies (*12*). The patients enrolled were extremely well characterized with four sputum cultures on two independent samples, blood cultures, large volume urine Xpert MTB/RIF and 2-month follow-up to ensure resolution of symptoms in the absence of treatment in patients who had a negative microbiological work-up. This extensive characterization allowed for a very good estimate of the true TB status of patients, which is typically extremely difficult in PLHIV (*13*). Additional analyses are currently under way using the Xpert-MTB-HR prototype from patients in diverse geographical locations to further evaluate its performance.

In summary, in this first accuracy study of a novel blood-based host-marker assay, we show the possible value of the assay for diagnosis in a vulnerable and very difficult to diagnose population living with HIV.

## Materials and Methods

### Study design and participants

We assessed biobanked blood samples collected as part of the FIND biobanking effort (https://www.finddx.org/specimen-bank/) from inpatients (>18 years) living with HIV, independent of CD4 count, in a South African district hospital and a Peruvian referral hospital between February 2016 and August 2017. Participants were adults with TB symptoms, able to produce sputum. Participants with presumed only extra-pulmonary disease were excluded. The samples were preserved in temperature-controlled freezers at -80°C from collection until testing.

The study was a nested case control study. Criteria for the selection of a sample were availability of PAXgene tubes (Becton Dickinson, Franklin Lakes, USA), in whom a comprehensive work-up was performed to identify TB or rule it out. Participants selected for this study (201 total) were representative of the larger cohort (259 participants with HIV) in respect to distribution of TB cases and smear status among the TB cases. Given the exploratory nature of the study, no sample size calculations were performed. All study-related activities were approved by the Human Research Ethics Committee (HREC) of the University of Cape Town (UCT) and the Universidad Peruana Cayetano Heredia (UPCH). Written informed consent was obtained from participants, as per study protocol. Study participation did not affect standard of care. Reporting followed STARD guidelines (*14*).

### Index test

Retrospective testing of PAXgene-conserved blood samples with the index test was performed by Cepheid in January 2019. The index test, the early Xpert-MTB-HR-Prototype, evaluates mRNA levels of three differentially expressed genes (*GBP5, DUSP3* and *KLF2*) (*5*). For the testing, one aliquot per patient of 380 µL thawed PAXgene-blood was centrifuged. The supernatant was gently decanted and any excess liquid from the side of the tube was removed. The pellet was resuspended with a lysis buffer (Cepheid) and vortexed. The lysate was then transferred to the Xpert-MTB-HR-Prototype cartridge and subsequently analyzed on a GeneXpert instrument. A TB-score was computed from Ct-values obtained by the GeneXpert analysis according to the formula suggested by Sweeney et al^8^: TB-score = [Ct*GBP5* + Ct*DUSP3*]/2 – Ct*KLF2*.

### Comparator test

Serum CRP levels were measured using the Multigent CRP Vario assay on Abbott Architect C8000 at Quest Diagnostics (*15*). The Multigent CRP Vario assay is a latex immunoassay. An antigen-antibody reaction occurs between CRP in the sample and anti-CRP antibody on latex particles. The resulting agglutination is detected as an absorbance change (572 nm), with the rate of the change being proportional to the quantity of CRP in the sample. The actual concentration is then determined by interpolation from a calibration curve prepared from calibrators of known concentration. For testing, in brief, the serum was thawed and then processed and interpreted following the manufacturer’s protocol.

### Reference standard

Fresh sputum, blood and urine specimens were processed using standardized protocols in centralized accredited laboratories of the South African National Health Laboratory Service and the UPCH. The testing flow for the samples tested are shown in the appendix (Supplement Figure 1). Standard testing was performed on all available sputum specimens and included Xpert MTB/RIF (Cepheid, Sunnyvale, CA, USA; testing predates rollout of Xpert MTB/RIF Ultra), smear fluorescence microscopy after Auramine staining, MGIT liquid culture (Becton Dickinson, Franklin Lakes, NJ, USA) and solid culture on Löwenstein-Jensen (LJ) medium. The presence of *Mycobacterium tuberculosis (M*.*tb)* complex in solid and liquid culture was confirmed with MPT64 antigen detection and/or the MTBDRplus line probe assays (Hain Lifesciences, Nehren, Germany). Blood cultures from all participants were done in BACTEC(tm) Myco/F Lytic culture vials (Becton Dickinson, Franklin Lakes, NJ, USA) and WHO-prequalified in-vitro diagnostic tests were used for HIV testing (rapid diagnostic tests) and CD4 cell counting (flow cytometry). For urinary Xpert MTB/RIF testing, 20-40 ml urine was centrifuged and following removal of supernatant, the pellet was re-suspended in the residual urine volume and 0.75 ml was tested using Xpert. The operators of the index were not blinded to the results of the reference standard.

### Case definitions

Participants were assigned to diagnostic categories using a combination of laboratory and clinical findings (Supplement Table 1). This was done by clinical investigators prior to performing the index test. Using a comprehensive microbiological reference standard (CMRS), “Definite TB” included participants with microbiologically confirmed *M*.*tb* (any culture from sputum or blood or any Xpert MTB/RIF from sputum or urine positive for *M*.*tb* during admission). “Not-TB” were participants with all microscopy, cultures and Xpert MTB/RIF tests negative for *M*.*tb* (and at least one non-contaminated culture result), who were not started on anti-TB treatment and were alive and improved at two months follow-up. “Not-TB” was further subdivided into patients with or without evidence of latent TB infection based on testing with commercial Interferon-Gamma-Release-Assays (IGRA). “Possible TB” included any patient not meeting “Definite TB” or “Not-TB” classification who is started on TB treatment. “Subclinical TB” was defined as patients presenting with clinical symptoms suggestive of TB, negative microbiological work-up at the time of first presentation but positive microbiological work-up at follow-up two months after first presentation. Participants with “Subclinical TB” and “Possible TB” were excluded from the primary analysis and reported separately (n=5).

In a separate analysis, “Definite TB” was defined by Xpert MTB/RIF positivity on the first sputum only (“Xpert MTB/RIF only” reference standard). This reference standard was used to most closely mimic what is likely to be the confirmatory test in limited resource settings (as recommended in the TPP by WHO and based on the fact that sputum culture has seen limited penetration in limited resource settings due to infrastructure needs and slow turn-around time) (*1*). “Not-TB” in this analysis were all participants who were Xpert MTB/RIF test negative on first sputum.

### Analysis

All samples had complete data on the reference standard testing. The diagnostic performance of the obtained TB-score was evaluated with a ROC (Receiver Operating Characteristic) curve analysis using the diagnostic categories of “Definite TB” and “Not-TB” as binary reference standard for the classifier. In the primary analysis, the categories “Definite TB” and “Not-TB” were defined by the CMRS. In a secondary analysis, the categories “Definite TB” and “Not-TB” were defined based on the Xpert MTB/RIF result on the first sputum as a reference standard.

Three separate analyses were performed for each of the two different reference standards:

1.) Sensitivity and specificity were calculated at the threshold value that maximized the Youden index

2.) In addition, to evaluate if the test fulfills the WHO requirements for a triage test, specificity was calculated at the closest threshold value corresponding to 90% sensitivity (minimal target)

3.) The same analysis as in 2.) was performed for a sensitivity of 95% (optimal target).

In addition, we explored if Xpert-MTB-HR-Prototype performance would meet the TPP for a non-sputum-based stand-alone diagnostic test, where specificity is optimized (*1*). While the specificity was set by WHO at 98% in the TPP, we decided to set it at 95% in our analysis as we recognize the limitations of the reference standard particularly in PLHIV. We restricted this analysis to a comparison against a CMRS only. Confidence intervals reported were calculated based on the Wilson method.

In the statistical analysis plan, the following sub-groups were prespecified for a sub-group analysis:

- smear status
- CD4 count (categorized as ≤200 and >200)
- number of symptoms (categorized as ≤3 or >3)

Symptoms were specified as cough, fever, night sweats, weight loss for at least two weeks. Another prespecified analysis included an assessment of specificity in the two subgroups of “Non-TB” without LTBI and “Non-TB” with LTBI based on IGRA results. Indeterminate results were not included in the analysis but reported separately.

## Data Availability

None

## Acknowledgments

The authors thank Audrey Albertini and Ranald Sutherland for helping with the conceptualization of this work, the participants who provided the samples and the clinical and laboratory teams at the partner sites for their efforts in performing the study.

## Author contributions

ES, DHP, SB, LSM, JP, and PK contributed to test design and prototype development SO, AM, RW, PK, and DG contributed to data analysis and evaluation. SGS and CMD contributed to clinical study design and data analysis.

---

**Supplement Figure 1:**
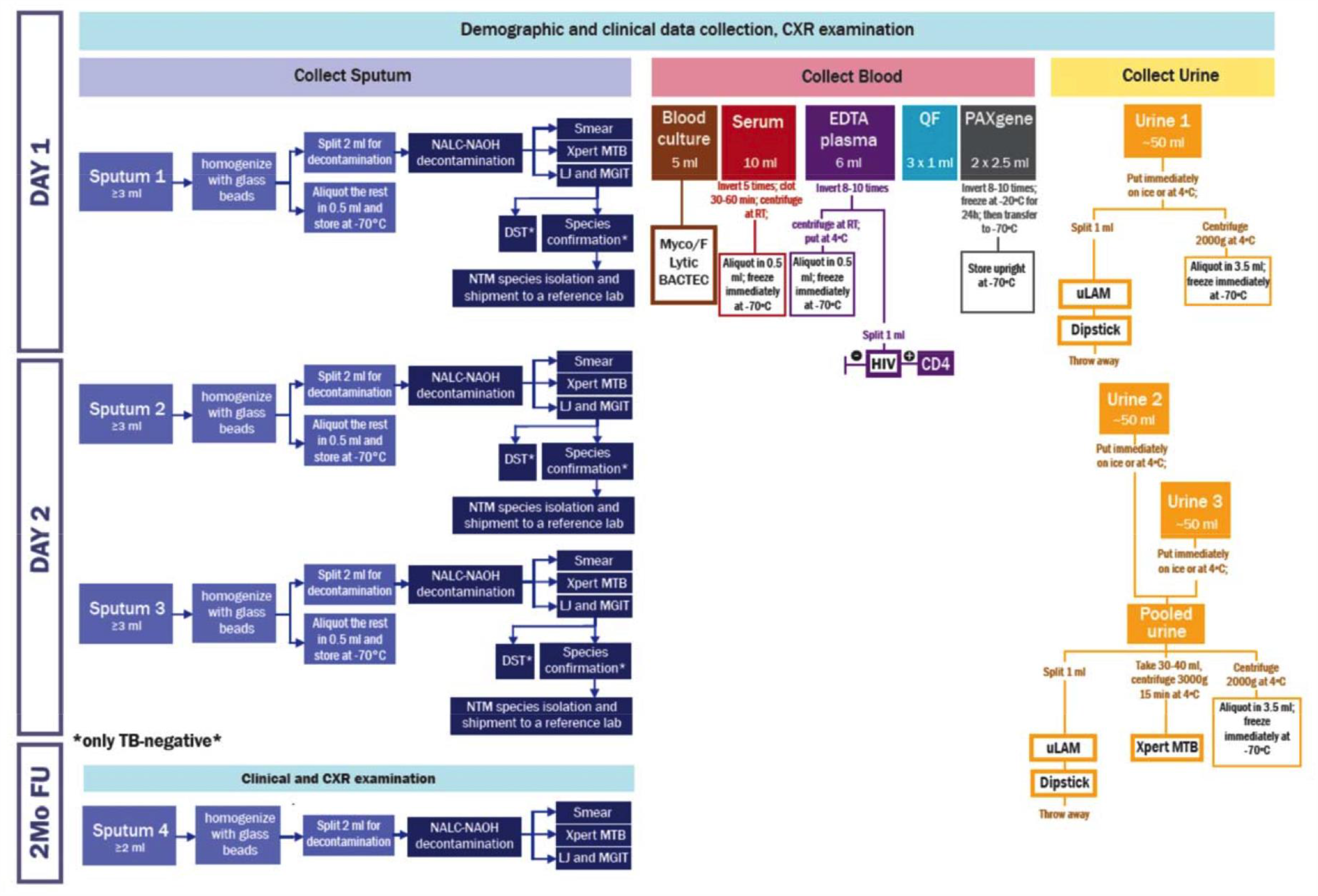
Specimen collection flow.

**Supplement Figure 2:**
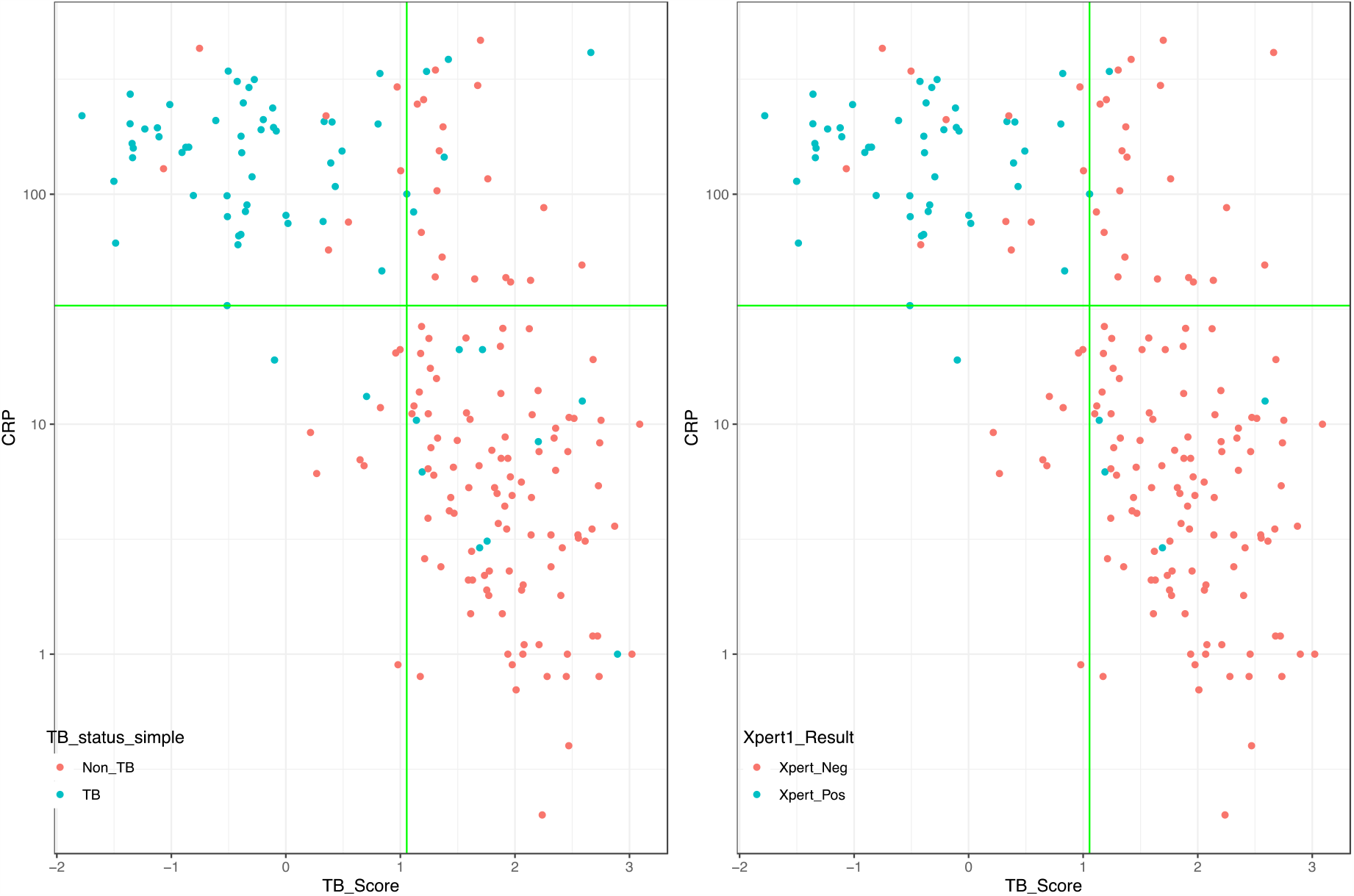
Correlation of CRP and TB-score of Xpert Prototype. CRP on Y-axis in log10-scale. TB-score on X-axis. The vertical and horizontal green lines represent 90% sensitivity threshold values for TB-score and CRP (29.7 mg/L). In case of CRP, samples detected at the 90% sensitivity threshold values will be the ones above the horizontal green line; for TB score, those will be the ones on the left of the vertical line. Color indicates which of those samples will be positive by the Xpert on the first sputum. Dots in the left panel are colored based on CMRS results; right panel colored based on results from Xpert on first sputum. When comparing Xpert Prototype and CRP scores for each sample, we observed that the nine samples mis-classified by TB-score and CRP are largely the same (see 4th quadrant). Xpert on sputum also does not identify 5 out of these 9 patients identified by CMRS. Furthermore, this figure shows that when using Xpert alone on first sputum as a confirmatory test, the Xpert Prototype has a smaller number of false positives (20 FPs) compared to CRP (31 FPs), which increases the specificity of Xpert Prototype, and result in lower number of Xpert cartridges required as a confirmatory test compared to CRP.

**Supplement Figure 3:**
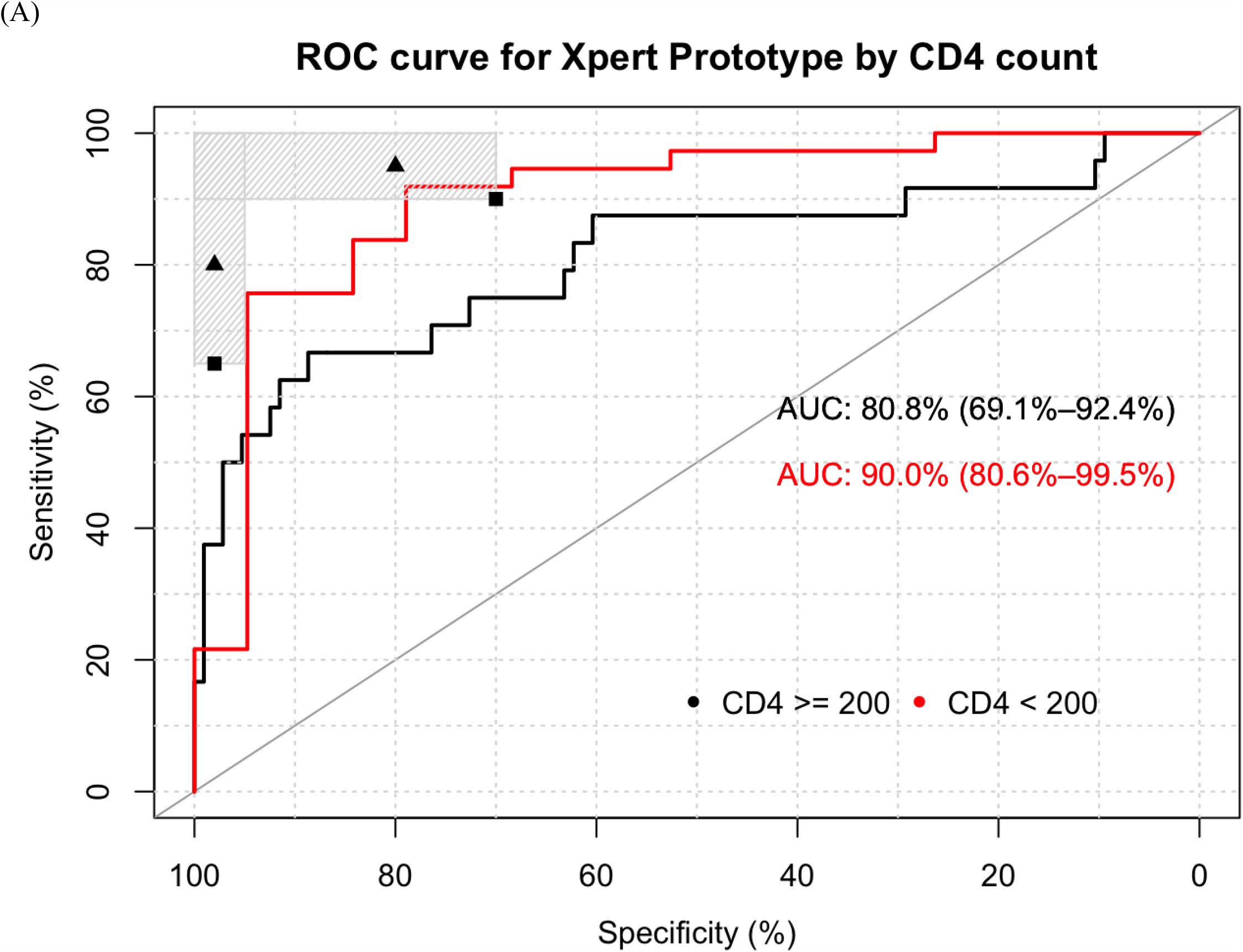
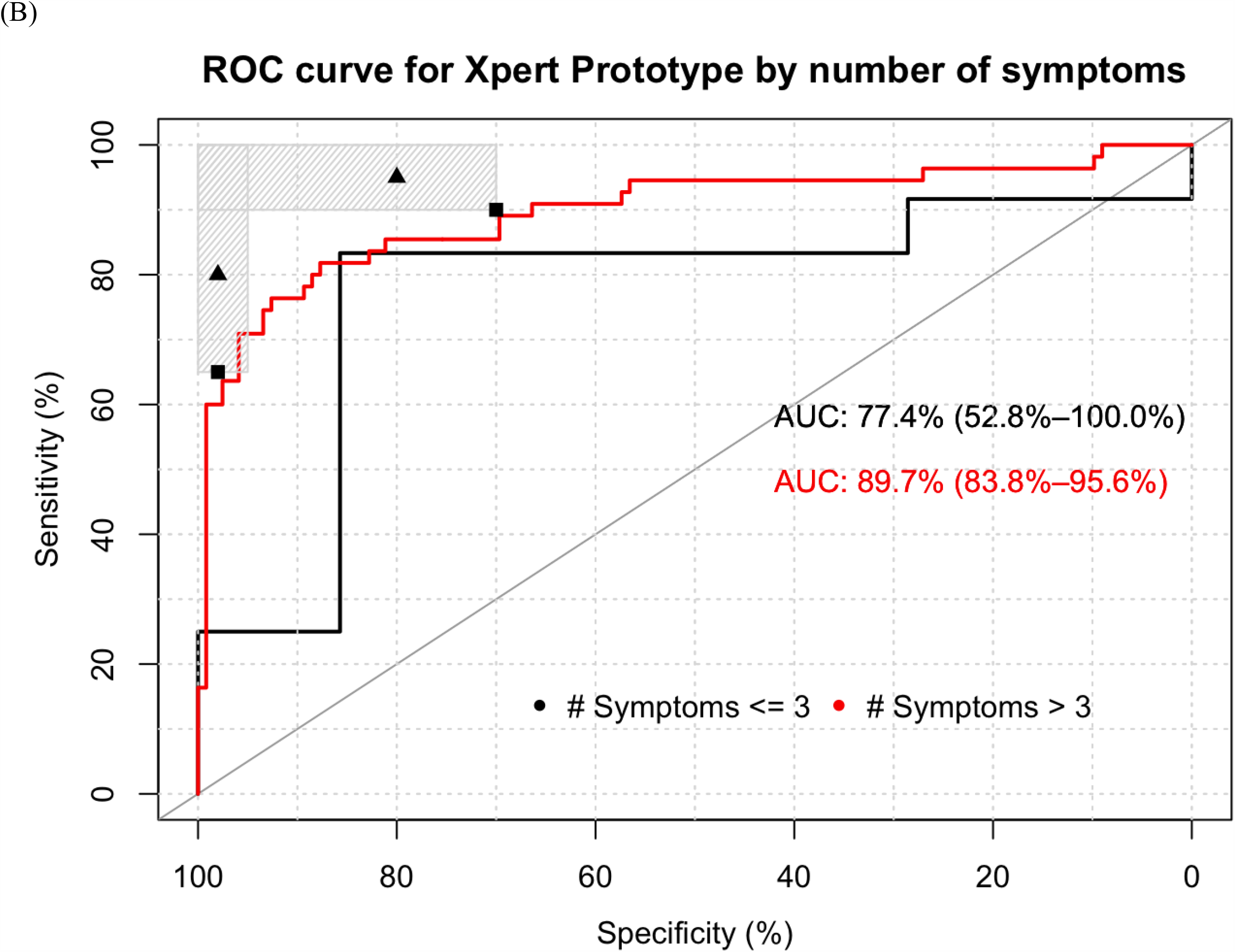
ROC curve for subgroups analyses (A) By CD4 count and (B) for number of symptoms at optimal TB-score cut-point against Xpert on first sample as reference standard.

**Supplement Figure 4:**
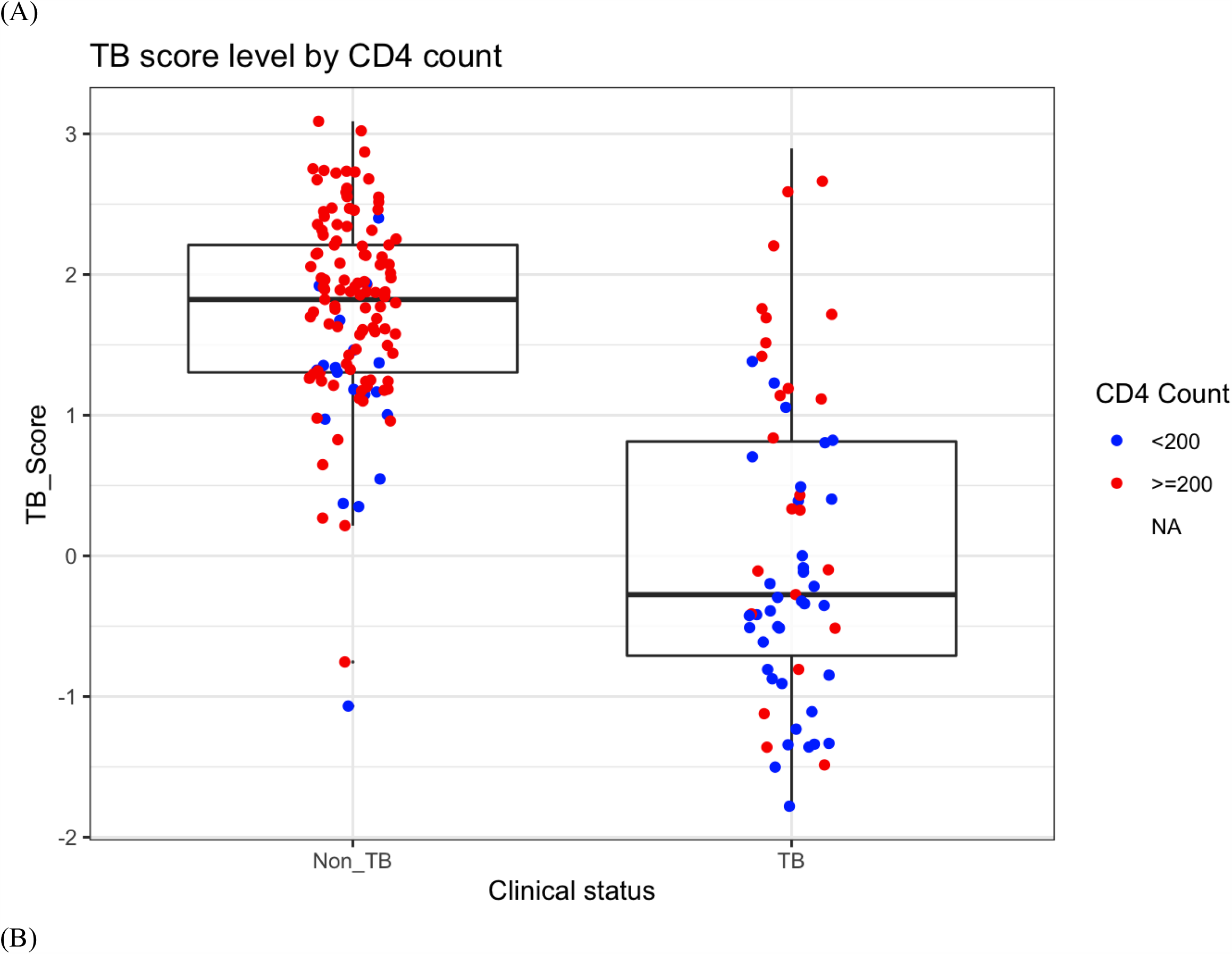
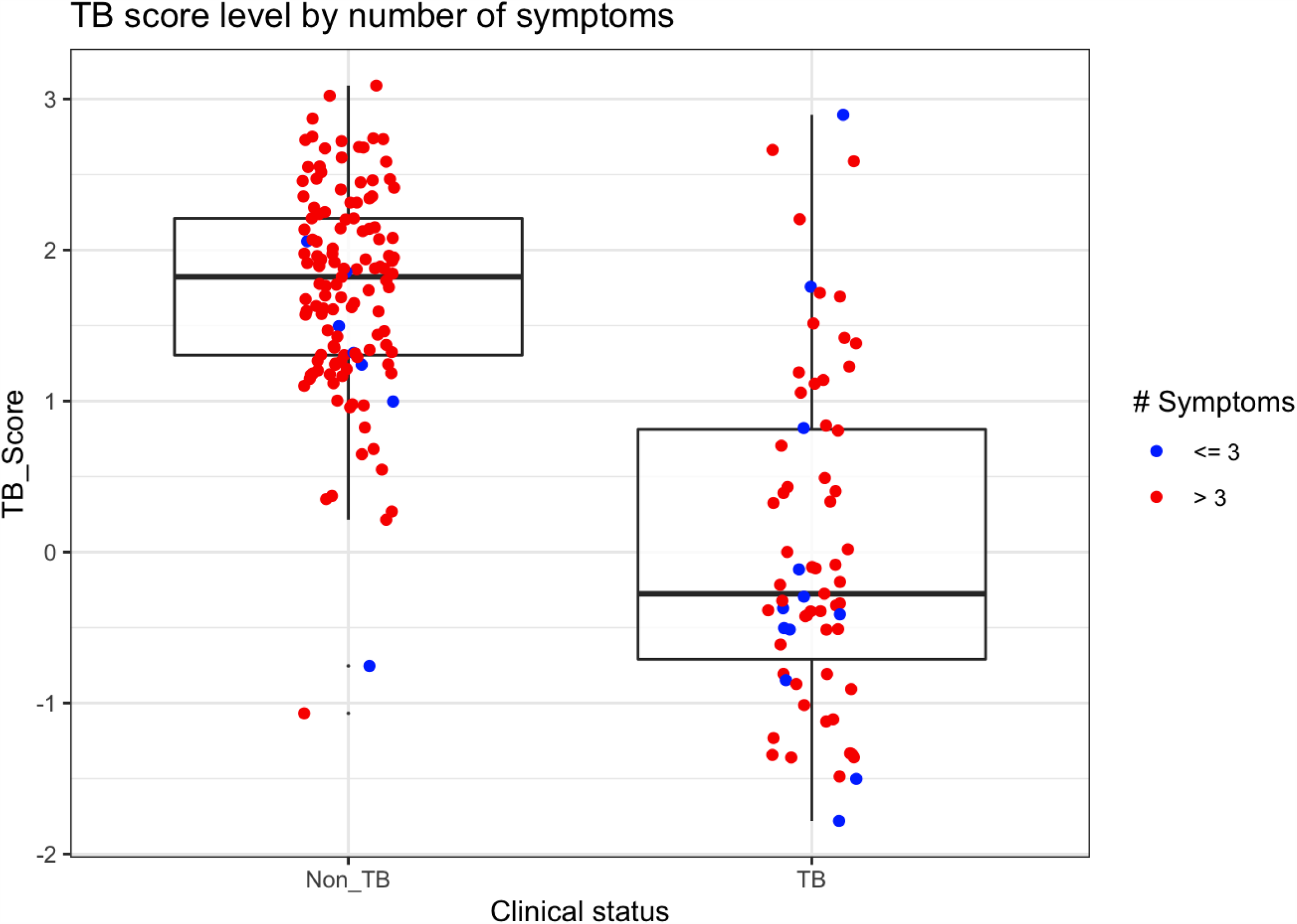
Box plots for subgroups analyses (A) By CD4 count and (B) for number of symptoms at optimal TB-score cut-point against Xpert on first sample as reference standard.

**Supplement Figure 5:**
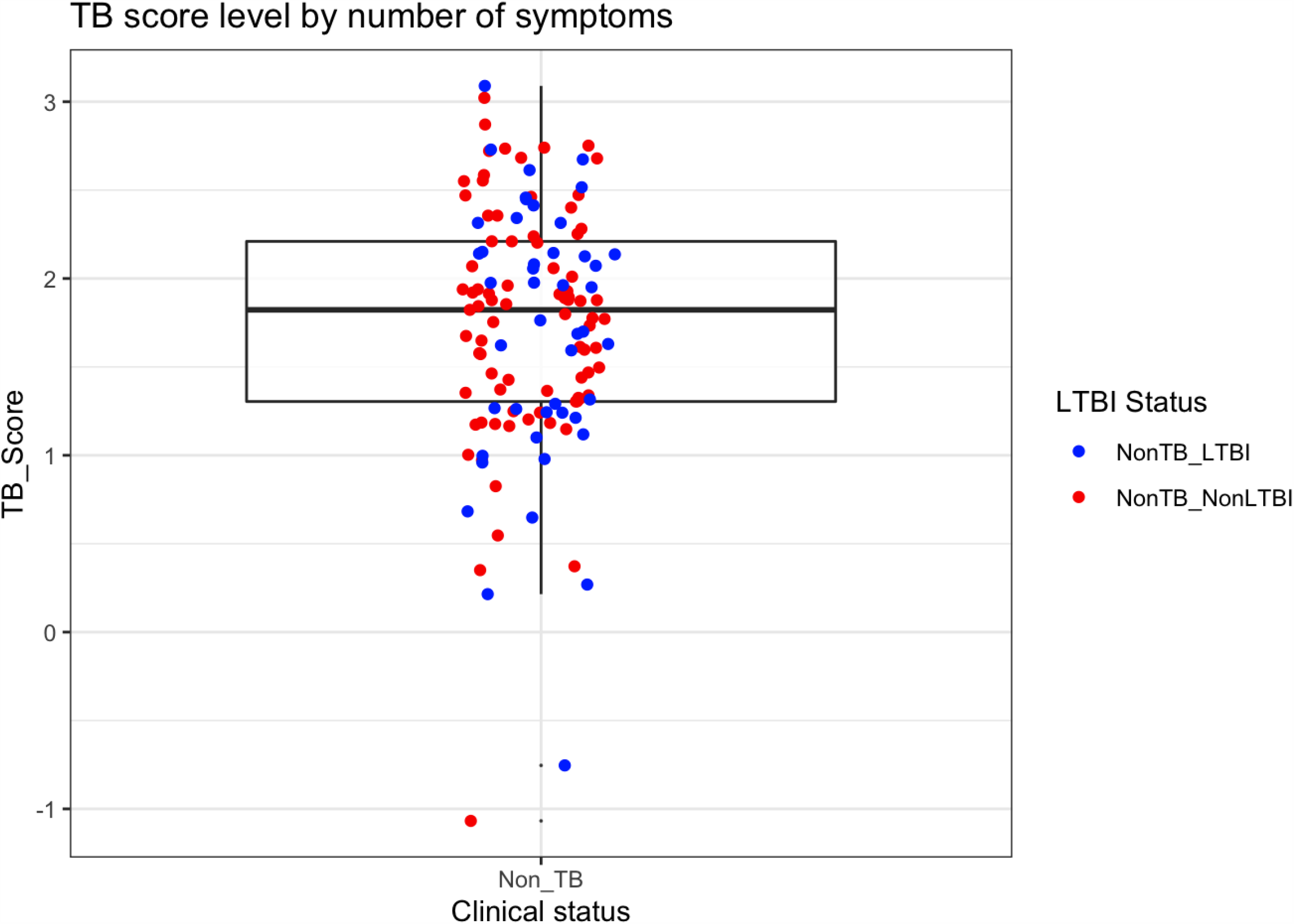
Box plots by LTBI status.

**Supplement Figure 6:**
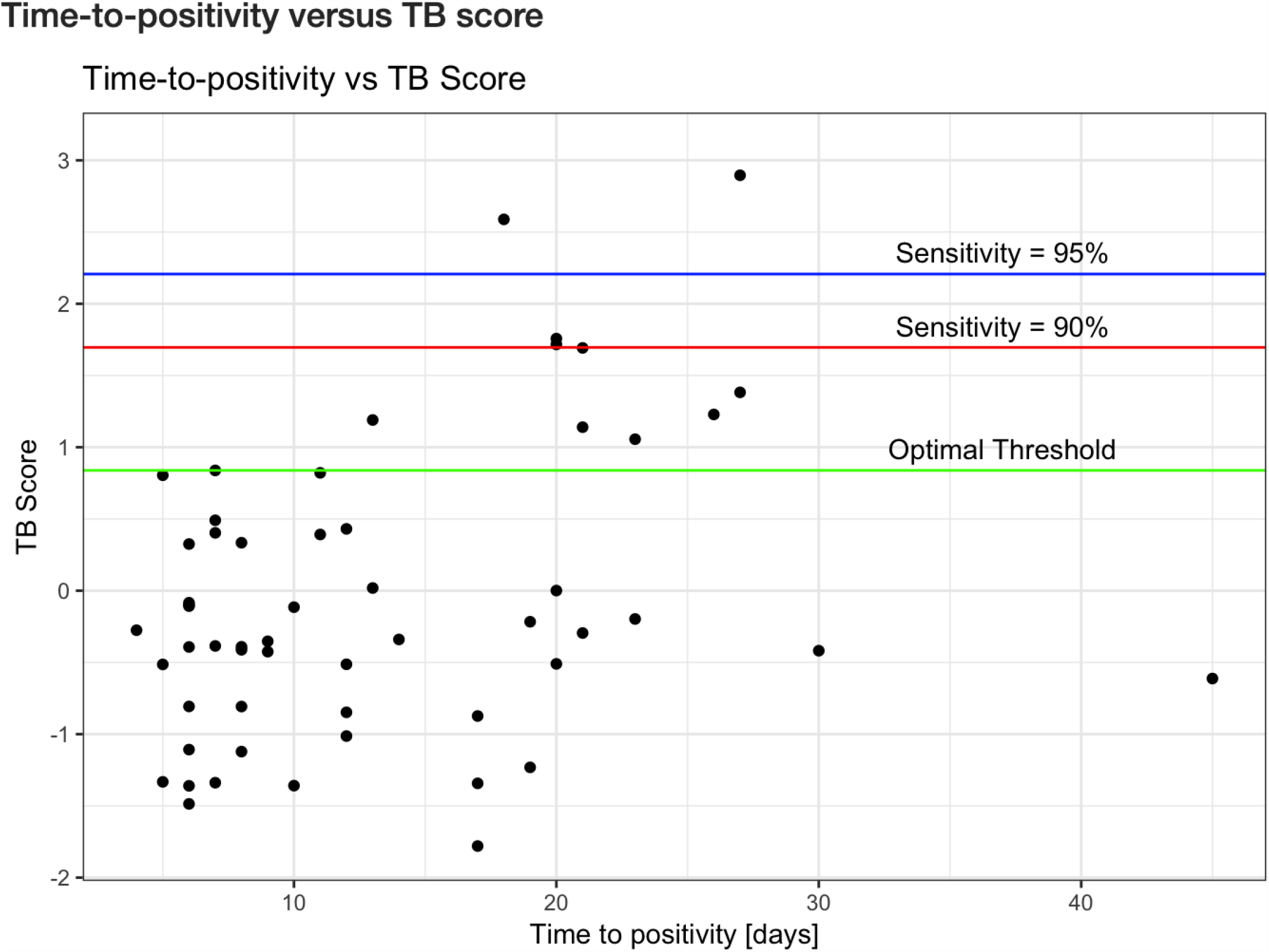
Time to positivity on culture versus TB-score of Xpert Prototype. Caption: Green line at TB-score cut-point with an optimized AUC; Red line: TB-score cut-point set to the minimal sensitivity of a triage test at 90%; Blue line: TB-score cut-point set close to the optimal sensitivity of a triage test at 95%.

**Supplement Table 1:**
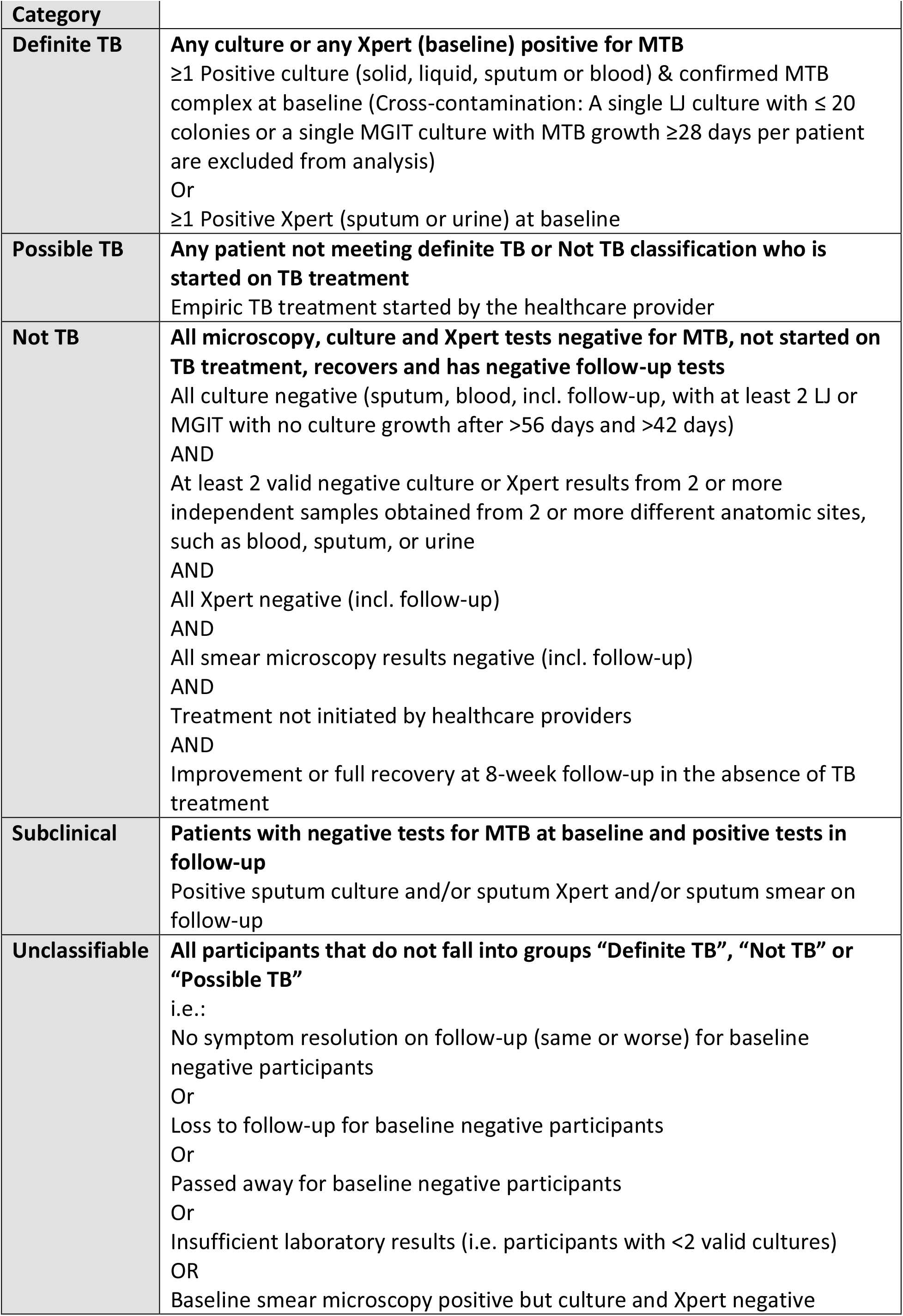
Diagnostic categories.

